# What is the value of community oximetry monitoring in people with SARS-CoV-2? – A prospective, open-label clinical study

**DOI:** 10.1101/2021.01.03.21249168

**Authors:** Jane Wilcock, Ciaran Grafton-Clarke, Tessa Coulson

## Abstract

**Background:** In people with COVID-19, hypoxia at the time of admission is known to be related to mortality. Monitoring of oxygen saturations (SpO_2_) is therefore an increasingly common part of community-based care, with the aim of improving the identification of adults who are deteriorating. We set out to investigate whether rigid SpO_2_ triggers, or absolute change in SpO_2_, is more indicative of deterioration in COVID-19.

**Methods:** A prospective, uncontrolled, open-label study in a large UK general practice was conducted between May and November 2020. Participants recorded twice daily oximetry and symptom diary for 14 days after test-confirmed COVID-19. Primary outcomes were the proportion of people whose SpO_2_ dropped to ≤ 94% and ≤ 92%, the average maximum reduction in SpO_2_, and admission to hospital. We also investigated the relationship between MRC Dyspnoea scale, modified Roth score, and SpO_2_ through correlation analyses.

**Results:** 52 participants were recruited, following which 41 participants completed the study. The average age was 45.9 years with 63.4% identifying as female. The mean maximum reduction in SpO_2_ was 2.8%. The average time to maximum reduction in SpO_2_ was 6.4 days. Nine participants (22.0%) had a reduction in SpO_2_ to ≤94%. Three of these had a reduction in SpO_2_ to ≤92%, for which all three were admitted to hospital. Modified Roth score and SpO_2_ were weakly positively correlated (.31). MRC dyspnoea scale score and SpO_2_ were moderately negatively correlated (-.53).

**Conclusions:** A reduction in SpO_2_ to ≤92% was found to be highly predictive for admission to hospital. Modified Roth score or MRC dyspnoea scale scores should not be used as proxy measures for oximetry. This study contributes to the ongoing narrative around community-based oximetry and provides insight and recommendations for those currently engaging in or planning to roll out similar schemes.

**Strengths and limitations of this study:**

- This study is pragmatically designed to answer an important clinical question in primary care.
- This study focused on previously published values of SpO_2_ for triggering escalation of care and therefore provides answers based on current clinical practice.
- 11 of the 52 patients who were recruited into the study did not return their oximeter or oximetry diary at the end of the study period.
- We did not validate the accuracy or reliability of the oximetry / symptom diaries, as these were self-completed by the participants themselves.
- Other than admission to hospital and mortality within the study period, no other clinical outcomes have been recorded.

**Funding statement:** This research received no specific grant from any funding agency in the public, commercial, or not-for-profit sectors.

**Competing interests:** Jane Wilcock has no competing interests to declare.

Ciaran Grafton-Clarke has no competing interests to declare.

Tessa Coulson has no competing interests to declare.

## Background

The COVID-19 pandemic has garnered the attention of the world. The United Kingdom has been markedly affected, with a devastating loss of life, impositions upon most aspects of life, and a perpetuating threat to our safety and welfare.^1^ Despite the herald of mass vaccination in suppressing community transmission and the hopeful trajectory towards the resumption of normalcy, the continued effort of the healthcare community in protecting the public remains an unwavering priority.^2^ As we surpass the sobering anniversary of COVID-19’s index case, we should take stock of our experiences and apply our learned wisdom in curtailing further loss of life.

An array of self-delivered assessment tools have been considered to alert patients and their care-providers of the earliest features suggestive of clinical deterioration.^34^ Given the inherent creativity and resourcefulness of the healthcare profession, it is unsurprising old and familiar tools have risen to the forefront of our battle against SARS-CoV-2. The oximeter is frequently hailed as the ‘fifth vital sign’, with its photoplethysmographic ability to non-invasively estimate the saturation of arterial blood with oxygen, underpinning its clinical utility across the breadth of medicine.^5^ Since hypoxia at the time of admission to hospital has been demonstrated to be a highly predictive determinator for mortality, it appears judicious to evaluate the utility of oximetry in identifying patients before they cascade into critical illness.^6^

The investment made by NHS England in procuring 200,000 oximetry devices for community use is laudable in ongoing efforts to protect vulnerable groups of patients.^7^ However, oximetry delivered without consideration of its integration into frameworks of community-based care, and determination of the quantitative triggers for escalation and intervention threatens potential benefits afforded by its use. There is an unmet clinical need for the academic community to evaluate the clinical usefulness of community oximetry monitoring.^8^ Clarity is required around whether rigid SpO_2_ triggers are appropriate, or instead, rate of change or absolute change is more indicative of the deteriorating patient.^8^ Providers need confidence that the delivery of monitoring in this way is sufficiently sensitive in identifying which people require medical attention, and at what time.

We sought to answer these crucial questions within this pragmatic research study by delivering oximetry monitoring alongside a symptom diary to over 50 adult patients with confirmed COVID-19. Here, we contribute to the ongoing narrative around community-delivered oximetry, whereby we provide valuable insight and recommendations for those currently engaging in similar schemes of practice or considering roll-out in their communities.

## Methods

### Study design and setting

We conducted a prospective, uncontrolled, open-label study from a large General Practice in the North-West of England. The study period ran from 15^th^ May to 30^th^ November 2020.

### Study population

Adults were eligible for inclusion if they returned a positive result for COVID-19 using reverse transcriptase-polymerase chain reaction (RT-PCR) of an oropharyngeal or nasopharyngeal swab and had symptoms. Patients were eligible for inclusion if they had been diagnosed with COVID-19 within the preceding seven days, were self-isolating in their residence, and were not living within a care or nursing facility.

### Study protocol

Once the results of COVID-19 tests had been released to the practice, patients were contacted via text message as an invitation for participation. Once enrolled, study packs were delivered to participants, which contained a CE approved oximeter, instructions for use, and a 14-day symptom and oximetry diary. Participants were instructed to measure their SpO_2_ using the oximeter twice per day, at the same 12-hour interval throughout. The symptom diary incorporated a spectrum of clinical symptoms commonly associated with COVID-19, including cough, temperature, and loss of smell. Participants were also asked to record the degree of breathlessness on the MRC dyspnoea scale and to record a Roth score.^9^ The Roth score as an assessment tool for acute breathlessness has been widely criticised due to lack of validation.^10^ We incorporated a modified Roth score into the symptom diary, which negated the requirement for unwell patients to time their expiration. Instead, participants were asked to count from 1 – 30, and to record the value they were able to phonate during a single exhalation. We aimed to correlate the modified Roth score with oxygen saturation values, to identify whether it has utility as a proxy for oxygen saturation in the absence of an oximeter. All participants were asked to report how useful they found the diary to be on a five-point Likert scale, and whether the diary caused anxiety, or relieved it. Participants were asked to complete 14-days of the diary or until complete recovery was achieved, whichever came first.

Within the information provided to participants, any drop in SpO_2_ equal to, or below 94%, participants were instructed to contact 111 for medical advice. If SpO_2_ dropped below 92%, or they felt they required urgent medical attention, then they were advised to call for the emergency services.

A small-group trial on six patients prior to study commencement highlighted several issues relating to oximeter reproducibility, which necessitated replacement by the manufacturer. This amounted to 14% (7/50) of the oximeters purchased. There were further issues around participants struggling to interpret the pulse oximeter interface, commonly misreading the reported values. Often, participants were mixing up the oximetry and pulse values. Therefore, the decision was made to mask the pulse reading on the device itself to reduce the risk of measurement bias. Additionally, the request for participants to record oximetry during exercise was removed to further simplify the recording of data.

### Outcomes

The primary outcomes were the proportion of patients whose oxygen saturation dropped ≤ 94%, the average maximum drop in oxygen saturation, the average time to maximum SpO_2_ drop, and the number of patients who were admitted to hospital. Secondary outcomes included the number of times healthcare services were accessed; proportion of time participants experienced cough, pyrexia, anosmia; correlation between modified Roth score, MRC dyspnoea score, and SpO_2_; diary completion rates; and user experience relating to anxiety and usefulness of the diary.

### Statistical analysis

Statistical analysis was performed using SPSS Statistics software version 27.^11^ Continuous variables are presented as mean ± standard deviation (SD). Pearson Product-Moment Correlation was used within the correlation analyses.

## Results

### Characteristics of study subjects

52 participants were recruited into the study (**Figure 1)**. Data is available for 41 participants. 11 recruited patients were not included in the data analysis. Four patients failed to return the oximeter and symptom diary. A further seven withdrew from the study for a variety of reasons, including family bereavement and anxiety over relatives who also had a diagnosis of COVID-19. One participant died (due to COVID-19) during the study, for which the symptom diary was not available for analysis.

**Figure 1:**
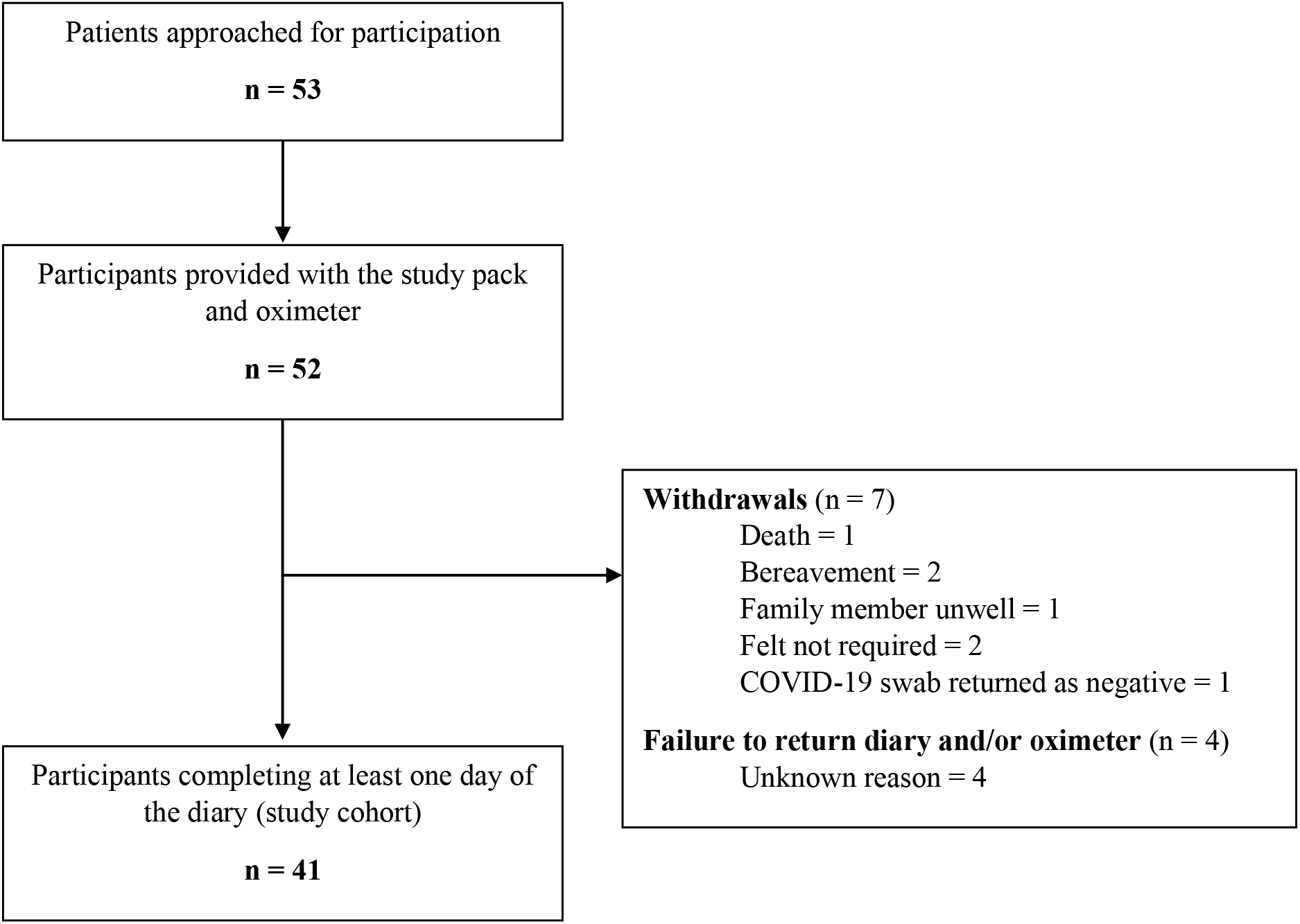
CONSORT diagram

Demographic and baseline characteristics of 41 participants is summarised in **Table 1**. The average age was 45.9 ± 8.7 years and the majority identified as female (63.4%). The level of comorbidity was low, corresponding to an average Charlson Comorbidity Index score of 1.2 ± 0.9. Of those returning the diary (n = 41), seven completed all 14 days (17.1%). Ten participants terminated completion of the diary before the end of the 14-day study period due to complete recovery of symptoms (17.1%). Of the 41 participants returning the diary, the average respondent completed 10.3 ± 1.4 days of it.

**Table 1:** Baseline characteristics, diary completion rates, outcomes, and symptomology for 41 participants

| **Baseline Characteristics** |  |
| --- | --- |
| Age (years) | 45.9 ± 8.7 |
| Sex (female) | 26 (63.4) |
| Charlson Comorbidity Index (CCI) | 1.2 ± 0.9 |
| CCI 0 | 25 (61.0) |
| CCI 1 | 4 (9.8) |
| CCI 2 | 1 (2.4) |
| CCI 3 | 5 (12.2) |
| CCI 4 - 7 | 6 (14.6) |
| **Diary Completion Rates** |  |
| Number of diary days completed | 10.3 ± 1.4 |
| Number of participants completing all 14 days | 7/41 (17.1) |
| Number terminating diary due to recovery | 10/41 (24.4) |
| **Patient Outcome** |  |
| Admission to hospital | 3 (7.3) |
| Mortality | 0 (0.0) |
| Recovered | 41 (100.0) |
| **Symptomology** |  |
| **Cough** |  |
| Cough for at least 1 diary day | 33 (80.5) |
| Number of days with cough (days) | 6.4 ± 1.7 |
| Proportion of diary days with cough (%) | 62.0 ± 14.5 |
| **Pyrexia** |  |
| Pyrexia for at least 1 diary day | 12 (30.8) |
| Number of days with pyrexia (days) | 3.0 ± 1.2 |
| Proportion of diary days with pyrexia (%) | 29.3 ± 12.0 |
| **Anosmia** |  |
| Anosmia for at least 1 diary day | 24 (58.5) |
| Number of days with anosmia (days) | 5.7 ± 1.8 |
| Proportion of diary days with anosmia (%) | 55.1 ± 14.1 |

*Values are mean ± SD or n (%).*

### Oximetry results

Mean maximum reduction in SpO_2_ was 2.8 ± 0.8%. The average time to maximum drop in SpO_2_ from the time of diagnosis was 6.4 ± 1.5 days. The reduction in SpO_2_ ranged from 1 – 7%. Nine participants (22.0%) had a reduction in SpO_2_ to at least 94%. Of the nine participants, three of these patients dropped their oxygen saturations to 92% or below (7.3%). All three of these patients were admitted to hospital. Of the six participants dropping their oxygen saturations to 93% or 94%, none were admitted to hospital. Two of these six participants were reviewed by the COVID-19 team, but no further action was felt necessary. The remaining four participants did not access healthcare services. The maximum and minimum oxygen saturations for these nine patients is presented in **Figure 2**. The trend in oxygen saturations of the five participants requiring intervention throughout the study period is presented in **Figure 3**.

**Figure 2:**
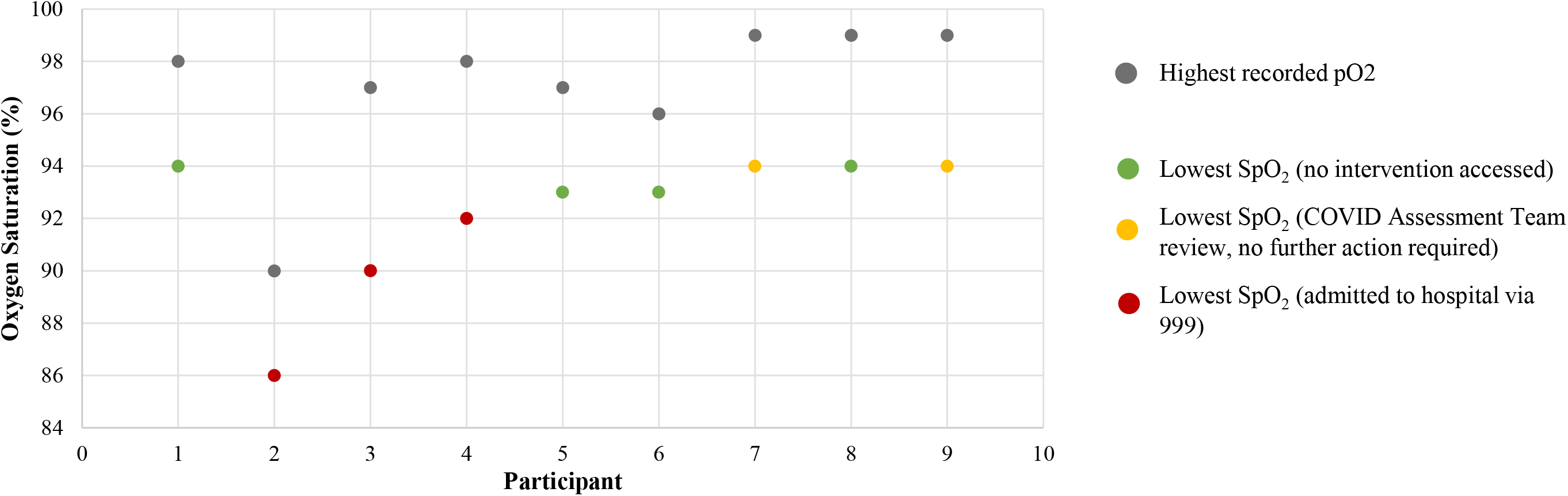
Maximum and minimum oxygen saturation in nine participants dropping their SpO_2_ to at least 94%

**Figure 3:**
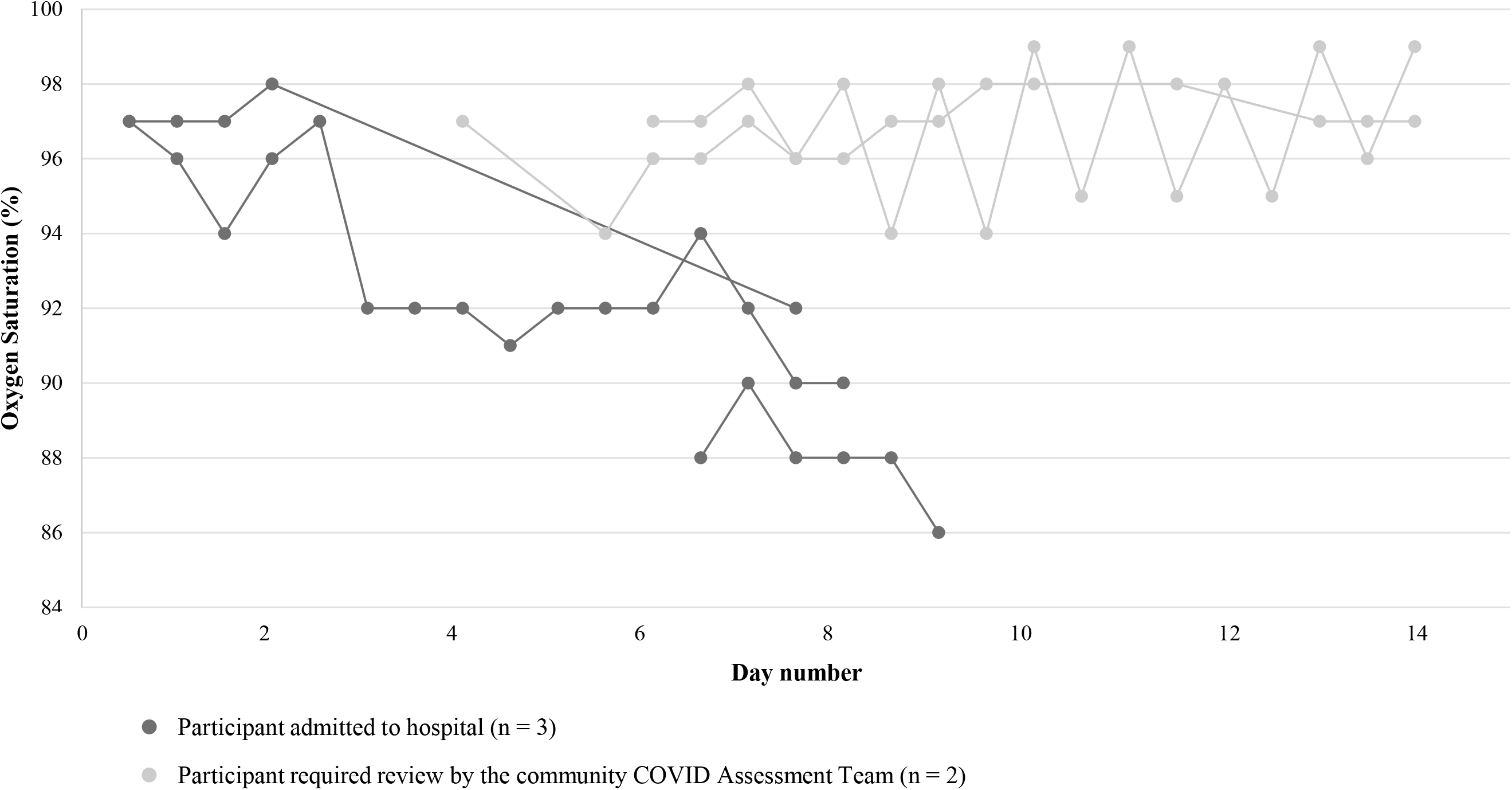
Oximetry trends in five patients for whom SpO_2_ dropped below 94% and health services were required

**Figure 4a:**
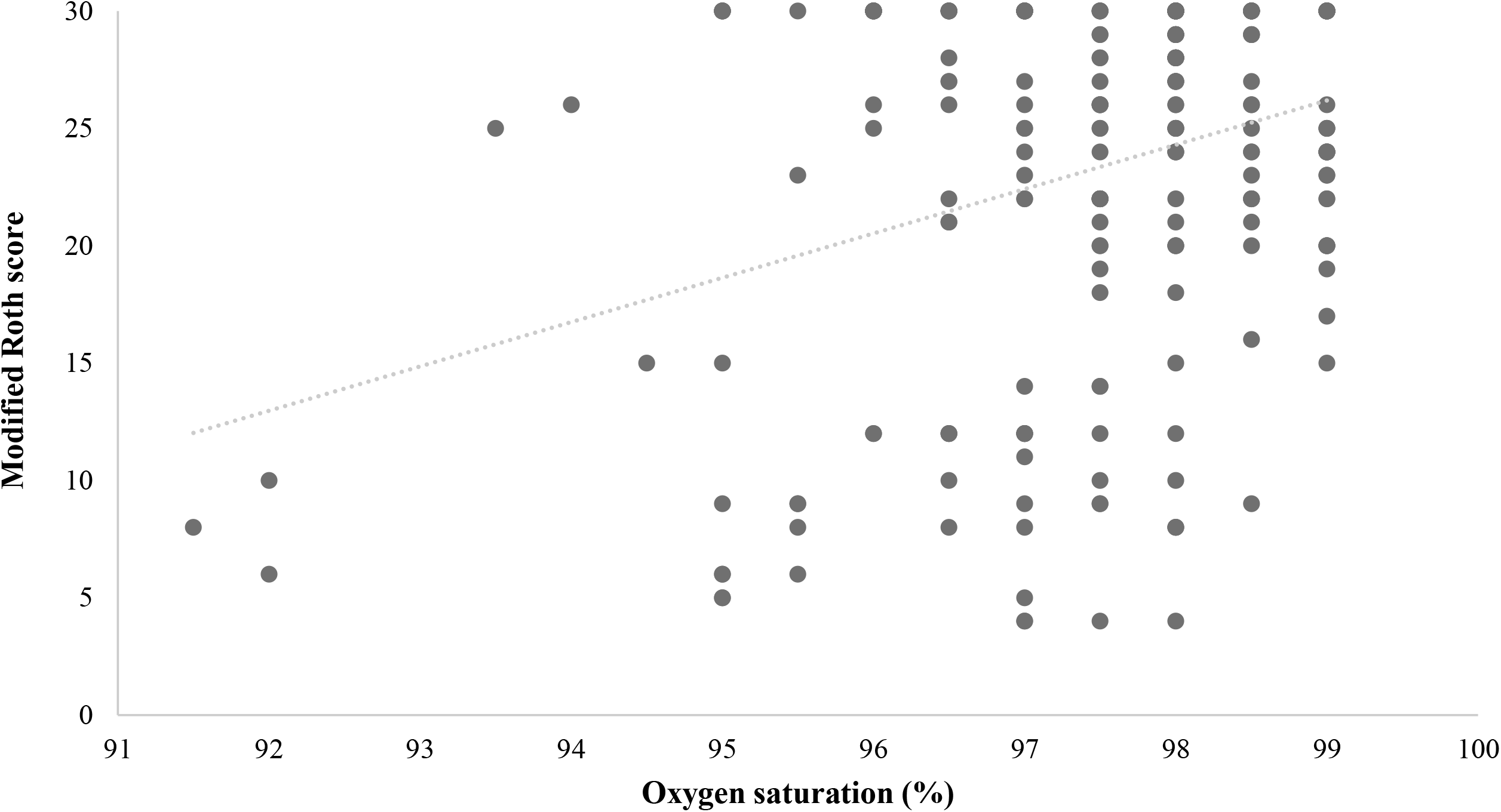
Correlation analysis between modified Roth score and oxygen saturation

**Figure 4b:**
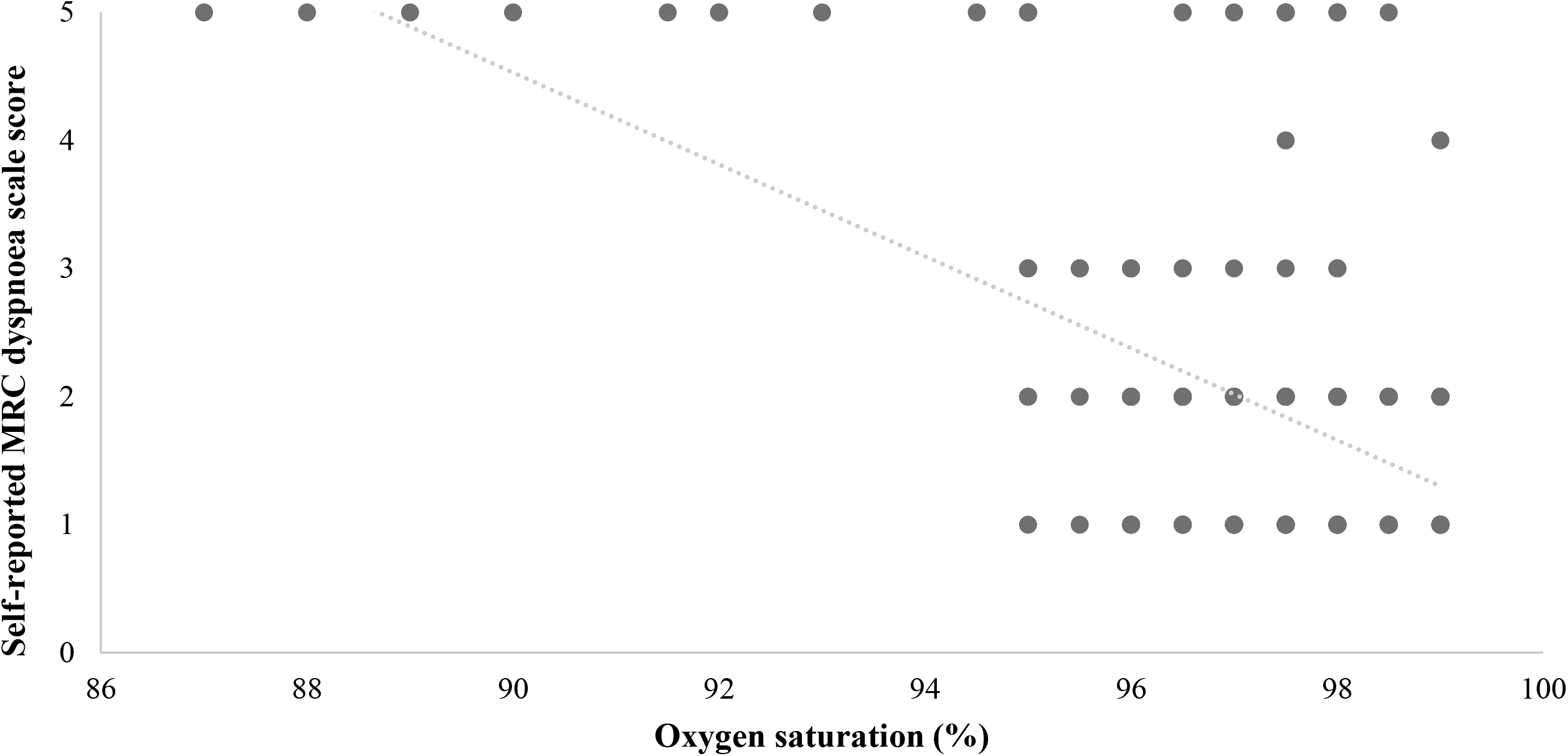
Correlation analysis between self-reported MRC dyspnoea scale score and oxygen saturation

In two of the three patients who were ultimately admitted to hospital, their oxygen saturation dropped by 5% and 7% respectively. The two patients requiring review by the COVID-19 team both dropped their oxygen saturations by 4%. Of the remaining 32 participants who dropped their oxygen saturations by no more than 3%, there was no requirement for COVID-19 team review or admission to hospital.

### Healthcare services accessed

Ten participants accessed health services through the study period, totalling 18 episodes of contact. The initial contact was with the GP for eight participants. In four cases, the GP referred the patient to the COVID Assessment Team for community review. Two participants contacted out-of-hours services. Four participants contacted the emergency services, following which three were transported to A&E for assessment and treatment.

### Symptom Diary

Cough, anosmia, and pyrexia were frequently described (**Table 1)**. Cough was a feature in 33 participants (84.6%). Of those recording cough as a feature, it was present for an average of 6.4 ± 1.7 days, equivalent to 62.0 ± 14.5% of the total diary days recorded. Anosmia affected 24 participants (58.5%) and was less sustained in comparison to cough, present for 5.7 ± 1.8 days, or 55.1 ± 14.1% of recorded diary days. Pyrexia was the least prevalent, affecting 12 participants (30.8%) for an average of 3.0 ± 1.2 days. 29.3 ± 12.0 of total diary days were affected by pyrexia. 76% of participants reported fatigue for at least one day within the study period.

### Modified Roth score and MRC dyspnoea scale

Modified Roth score and oxygen saturation were found to be weakly positively correlated, r (240) = .31 (**Figure 6**). There was a wide distribution of modified Roth scores for a given SpO_2_. For example, in participants with an SpO_2_ of 97%, the modified Roth score ranged from 7 to 30.

Self-reported MRC dyspnoea scale score and oxygen saturation were found to be moderately negatively correlated, r (288) = -.53. As with the modified Roth score, there was a wide distribution in MRC dyspnoea score for a given SpO_2_. For example, in participants with an SpO_2_ of 97%, self-reported MRC score ranged from 1 to 5.

### Participant satisfaction score

Using a five-point Likert scale (1 = much more anxious; 5 = much less anxious), participants reported the diary as reducing anxiety, corresponding to an average score of 4.0. Similarly, participants found the diary to be useful, reporting an average 5-point Likert score of 4.6 (1 = not at all useful; 5 = very useful).

## Discussion

In this study, we assessed the usefulness and value of home oximetry in monitoring patients with COVID-19. This study was pragmatically designed to identify whether the use of community-delivered oximetry monitoring would help identify the clinically deteriorating patient and whether it was acceptable to patients with COVID-19.

While we recognise the limitation around the relatively small sample size within this study, there are some interesting findings which should be explored further within large outcome-based clinical trials. For instance, we identified a drop in SpO_2_ to 92% to be highly predictive for hospital admission. This mirrors the conclusion drawn from a larger observational study delivered in COVID-19 patients following a discharge from A&E, where SpO_2_ values below 92% were predictive for re-hospitalisation.^12^ Our study has reinforced the cut-off value of 94% or 93%, as outlined by NHS England to be a trigger for assessment by a healthcare professional, and for an oxygen saturation of 92% or below to be a prompt for urgent admission to hospital.^6^ Three patients within our study cohort dropped their oxygen saturation to at least 92%. In all cases, it was the drop in oxygen saturation, accompanied by a deterioration in physical condition, which prompted a call to the emergency services, which in two cases was made by the next-of-kin, who incidentally had COVID-19 also. Whilst it is conjecture to speculate whether the oximetry monitoring conferred a mortality and morbidity advantage, it is reassuring that all those admitted to hospital were ultimately discharged from hospital. While the value that the oxygen saturation dropped to was an important indicator, the absolute reduction in SpO_2_ was also an important sensitiser towards clinical deterioration. For example, in two of the three patients who were ultimately admitted to hospital, the oxygen saturation dropped by more than 5%. Further evaluative research is required to determine whether absolute reductions or cut-off values of SpO_2_ are more sensitive indicators of deterioration necessitating medical intervention. We question whether widespread recording of resting oximetry in states of ‘health’ would be valuable for people who subsequently develop COVID-19, so that change from baseline could be measured.

Our recommendations for others who may be delivering similar schemes, which we predict may be sizeable given the drive from NHS England to utilise community oximetry to protect high-risk COVID-19 patients, to be as follows:

1. To offer oximetry for at least ten days following the diagnosis of COVID-19.
2. To encourage patients to contact 111 or their GP, if their oxygen saturations drop to 94% or 93%.
3. To inform patients to call 999 if their oxygen saturations drop to, or below 92%.
4. The Roth score and MRC dyspnoea scale should not be used as proxy measures for SpO_2_ or assessment of COVID-19 severity.

Participants within this study were offered oximetry monitoring for 14 days following the initial diagnosis of COVID-19, for which early completion was acceptable if recovery was achieved. Only a small number of participants performed oximetry for the entire 14 days, since a sizeable proportion stopped monitoring once they felt well in themselves. Several participants did not start monitoring their oxygen saturations until they felt their clinical condition was worsening, which in one case led to the identification of hypoxia with an SpO_2_ of 90% and falling. Given the six-day average interval between diagnosis of COVID-19 and maximum reduction in SpO_2_, we advocate that monitoring saturations for at least 10 – 12 days to be a safe approach for the majority of patients.

As with any monitoring test or tool, for which community oximetry should be considered as such, it must be acceptable to those who use it. Within this study, we have demonstrated that the participants placed a great deal of value in the comfort afforded by self-monitoring of oxygen saturations. Furthermore, the use of oximetry reduced levels of anxiety in the majority of participants, which is an important finding of this study, given the quintessential psychological burden a COVID-19 diagnosis imparts upon its sufferers. Reflecting further on the value of oximetry as a monitoring tool, there is good evidence highlighting the precision, safety, and accuracy of oximeters, provided they hold the relevant medical device approvals. Furthermore, there is increasing evidence, for which this study contributes to, that there is likely to be appropriate trigger values for escalation and intervention. Given the relatively low expense, re-usability, and ease at which oximeters can be decontaminated, there is very likely to be a significant opportunity cost for the use of community oximetry, provided it is an appropriately sensitive identifier of the deteriorating patients at an early stage. In doing so, healthcare professionals can go on to initiate established and evidence-based medical therapy.^13 14^

From our practical experience of delivering community oximetry monitoring, it is important for oximeters to be checked prior to use, and for patients to receive instruction to ensure data collection is accurate and undemanding. It was clear from the symptom diary that lethargy was a common feature, with the majority bedbound for a period of their illness. A common scenario was that oximetry was performed by worried relatives, many of them suffering with COVID-19 themselves. Decision-making in this situation can be difficult and reinforces the need to provide support for patients with COVID-19. In people living alongside those with COVID-19, the risk of co-infection from undertaking monitoring of their relatives is a burden.

We cannot advocate for the use of a modified Roth score or the MRC dyspnoea scale as proxy measures for oxygen saturations. Currently, there are no validated tests for the remote assessment of breathlessness in the acute primary care setting. We would therefore encourage primary care practitioners, in the absence of face-to-face consultation, to utilise video-consultation given the value of visual cues and the therapeutic presence, in addition to focused questioning on the velocity of change, impact of dyspnoea, and contextualisation within the wider illness.^15^

Given the potential for oximetry in protecting patients with COVID-19, there is a significant paucity of published research surrounding its application. Whilst this study provides some insights for further evaluation, there is a major need to appraise its use within larger cohorts of patients. For instance, the cohort of patients within this study were generally young and quite healthy. It therefore becomes challenging to apply the results of this cohort to significantly co-morbid populations, such as those who reside in nursing homes. We would strongly encourage any research groups or commissioning groups who are delivering similar modes of practice to evaluate its impact, and to particularly consider the sensitivity of trigger values, such as the ones we advocate for. We would also encourage those groups to relate community practice to clinical-outcomes, such as admission to intensive care, mortality rates, and long-term morbidity, preferably through a randomised-controlled trial.

## Data Availability

all data kept at the sponsors research site

## Contribution

Jane Wilcock was involved in study conception, study organisation, data collection, data analysis, writing of the manuscript, and approval of the submitted script.

Ciaran Grafton-Clarke was involved in data collection, data analysis, writing of the manuscript, and approval of the submitted script.

Tessa Coulson was involved in study conception, study organisation, data collection, data analysis, writing of the manuscript, and approval of the submitted script.

## Notes

### Competing Interest Statement

The authors have declared no competing interest.

### Clinical Trial

Independent UK GP research not registered.
HRA IRAS project ID: 283310 Ethics review: 20/HRA/2326

### Clinical Protocols

https://www.researchgate.net/publication/348170563_What_is_the_value_of_community_oximetry_monitoring_in_people_with_SARS-CoV-2_-_A_prospective_open-label_clinical_study

### Funding Statement

funded by chief investigator

### Author Declarations

HRA ethics committee approval 20/HRA/2326

